# Reactivation of Human Herpesvirus 6 and Epstein-Barr Virus in relapsing remitting multiple sclerosis: association with disabilities, disease progression, and inflammatory processes

**DOI:** 10.1101/2024.09.10.24313388

**Authors:** Abbas F. Almulla, Aristo Vojdani, Yingqian Zhang, Elroy Vojdani, Michael Maes

## Abstract

**Background:** Multiple sclerosis (MS) is a chronic autoimmune disorder affecting the central nervous system (CNS). Reactivation of Human herpesvirus 6 (HHV-6) and Epstein-Barr virus (EBV) is observed in MS.

**Objectives:** This study investigates immunoglobulins (Ig)G, IgM, and IgA directed against EBV nuclear antigen EBNA-366-406, HHV-6 and EBV deoxyuridine-triphosphatase (dUTPase), and different immune profiles in 58 patients with relapsing remitting MS (RRMS) compared to 60 healthy controls.

**Methods:** We employed enzyme-linked immunosorbent assays (ELISA) to measure the immunoglobulins to viral antigens. Multiplex immunoassays were used to measure cytokines, chemokines and growth factor levels that were used to compute immune profiles, including M1 macrophage, T helper (Th)-1, Th-17, and overall immune activation. We assessed disabilities using the Expanded Disability Status Scale (EDSS) and disease progression using the Multiple Sclerosis Severity Score (MSSS).

**Results:** IgG/IgA/IgM directed to the three viral antigens were significantly higher in RRMS than in controls. RRMS was significantly discriminated from controls by using IgG and IgM against HHV-6 dUTPase, yielding an accuracy of 91.5% (sensitivity=87.3% and specificity=95.2%). Neural network analysis showed that using IgG to EBV-dUTPase, IgM to EBV-dUTPase, and immune profiles yielded an area under the ROC curve of 1 and a predictive accuracy of 97.1%. There were strong associations between IgG/IgM responses to HHV-6 and EBV-dUTPases and the EDSS/MSSS scores and aberrations in M1, Th-17, profiles, and overall immune activation.

**Conclusions:** HHV-6 and EBV reactivation play a key role in RRMS and these effects are mediated by activation of cytokine profiles.

## Introduction

Multiple sclerosis (MS) is a chronic, autoimmune-mediated disease that primarily affects the central nervous system (CNS). MS is defined by the demyelination, neuroinflammation, and neurodegeneration of the nervous system ^1–3^, resulting in a wide range of symptoms, including impaired motor function, and sensory abnormalities ^4–6^. The global burden of MS is substantial, with approximately 2.8 million people affected worldwide, exhibiting significant geographical variability in prevalence ^7,8^. The manifestation of MS varies greatly, encompassing a wide range of severity levels and diverse clinical trajectories, such as relapsing-remitting MS (RRMS), primary-progressive MS, and secondary-progressive MS ^9^.

Recent research highlights the critical role of immune-inflammatory responses and oxidative stress in the onset of MS ^10–12^. Activated IRS pathways in individuals with MS are demonstrated by elevated concentrations of proinflammatory cytokines in both the blood and cerebrospinal fluid (CSF) ^13^. Even remitted RRMS patients show indicants of activation of the a) immune response system (IRS) with M1 macrophage, T helper 1 (Th1), T helper 17 (Th17) activation, b) compensatory immune response system (CIRS) with Th2 and T regulatory (Treg) cell activation, and c) chemokines and growth factor networks ^5^. Additionally, some studies found an association between the severity of clinical disabilities, as measured by the Expanded Disability Status Scale (EDSS) and indicators of oxidative and nitrosative stress ^14,15^.

The pathogenesis of MS involves the infiltration of immune cells such as T helper (Th)-17, B cells, and CD4+ and CD8+ T cells into the CNS ^16^. Furthermore, inflammation driven by T cells, B cells, macrophages, activated microglia, and reactive oxygen and nitrogen species play a key role in the loss of myelin and degeneration of the nervous system in all stages and forms of MS ^12,17^. However, the underlying mechanisms driving the activation of immune responses and the pathways leading to oxidative stress in MS remain incompletely understood.

Latent viral infections, particularly by Epstein-Barr virus (EBV) and human herpesvirus 6 (HHV-6), have been increasingly implicated in the pathogenesis of MS ^18^. These viruses have been detected in the CNS and CSF of MS patients, and their presence correlates with disease activity, suggesting a contributory role in disease onset and progression ^19–21^. The immunoglobulin (Ig) response to these viral proteins has been proposed as a marker for MS onset and progression ^22,23^. For example, elevated levels of IgM directed to HHV-6 (IgM-HHV-6) and IgG-EBV antibodies have been observed in early-stage MS patients ^24^. EBV nuclear antigen 1 (EBNA-1) immunoglobulin G (IgG) levels are associated with the disabilities observed in MS (as assessed with the EDSS) and with gadolinium-enhancing lesions ^25^.

However, the associations between IgM/IgA/IgG levels directed to HHV-6 and EBV deoxyuridine 5′-triphosphate nucleotidohydrolase (dUTPase) were not investigated in MS. The herpesvirus dUTPases are a reliable indicator of viral activity, as they are only expressed during viral replication and reactivation or active “abortive” infection ^26,27^. Moreover, as a pathogen-associated molecular pattern (PAMP), herpes dUTPases activate nuclear-factor κB and the host immune response ^28,29^.

Thus, this study examined whether a) the remitted phase of RRMS is accompanied by signs of EBV and HHV-6 replication or reactivation as assessed with IgG/IgA/IgM responses to EBV-and HHV-6-dUTPase and EBV nuclear antigen (EBNA-366-406); and b) whether the EBV or HHV-6 reactivation indices are associated with disabilities and progression of disease and the immune profiles observed in RRMS.

## Material and Methods

### Subjects

Between September 2021 and March 2022, 58 RRMS patients from the Neuroscience Center of Alsader Medical City in Al-Najaf province, Iraq, were enrolled in this case-control study. They were in the remitted phase of RRMS. The diagnosis of MS was confirmed using the McDonald criteria ^30^ by a senior neurologist. Additionally, sixty healthy individuals from the same demographic area, including hospital staff, their acquaintances, and medical workers were recruited.

All participants were screened to exclude those with current Axis-I neuropsychiatric disorders, such as schizophrenia, bipolar disorder, psycho-organic disorders, and substance use disorders (except nicotine dependence). The study also excluded individuals with medical conditions like diabetes mellitus, cardiovascular diseases, thyroid disorders, renal or liver diseases, gastrointestinal conditions, oncologic disorders, other (auto)immune, and neuroinflammatory and neurodegenerative diseases, including psoriasis, COPD, inflammatory bowel disease, Parkinson’s disease, and Alzheimer’s disease.

Written informed consent was obtained from all RRMS patients or their parents/legal guardians, as well as from control participants prior to their inclusion in the study. This research received ethical approval from the institutional ethics board of the College of Medical Technology at The Islamic University of Najaf, Iraq (document number 11/2021). Adherence to ethical standards was ensured by following both Iraqi and international guidelines, including the World Medical Association Declaration of Helsinki, The Belmont Report, CIOMS guidelines, and the International Conference on Harmonization of Good Clinical Practice (ICH-GCP). Our Institutional Review Board (IRB) complies with the International Guidelines for Human Research Safety.

### Clinical Assessment

A senior neurologist conducted a semi-structured interview to gather sociodemographic and clinical data, including duration of illness (DOI). The same neurologist utilized the EDSS ^31^ to evaluate clinical disability, and the Multiple Sclerosis Severity Score (MSSS) ^32^ to assess the progression of disability over time. Activities of daily living (ADL) were measured using the Arabic-translated index of ADL ^33^. Body mass index (BMI) was determined by dividing the participant’s weight in kilograms by their height in meters squared.

### Biomarkers assays

Blood samples were collected from fasting participants between 7:30 and 9:00 a.m. using venipuncture with disposable syringes. The collected blood was allowed to clot at room temperature for 15 minutes before being centrifuged at 3500 RPM for 10 minutes. The resulting serum was aliquoted into Eppendorf tubes for use in various assays. An enzyme-linked immunosorbent assay (ELISA) was employed to detect antibodies (IgA, IgG, IgM) specific to HHV-6-duTPase, EBV-DUTPase, and EBNA-366-406 peptides, which were synthesized from Biosynthesis (Lewisville, TX, USA). The methodology for these procedures is detailed in previous studies ^34,35^. In brief, 100 microliters of different peptides at a concentration of 5 micrograms per mL in 0.1 M carbonate buffer ph 9.5 were added to different wells of ELISA plates. After incubation, washing and blocking with 2% BSA, 100 microliters each of the control and MS patients’ sera at a dilution of 1:50 for IgA and 1:100 for IgG and IgM determination were added to duplicate wells. Plates were then incubated, washed, and secondary antibodies were then added to each plate. Finally, after repeated washing and the addition of substrate, the color development was measured, and indices were calculated using different sera as calibrators and controls.

Additionally, a comprehensive assessment of cytokines/chemokines/growth factors, relevant to M1 macrophage activation, Th-1, Th-17, IRS, CIRS, IRS+CIRS, chemokine, and growth factor profiles was performed using the Bio-Plex Pro™ Human Chemokine Assays from Bio-Rad Laboratories, Inc. (Hercules, USA) as described previously ^5^. The Electronic Supplementary File (ESF), Table 1, shows the cytokines/chemokines/growth factors assessed in this study. As described previously ^36,37^, our analysis utilized immunofluorescence (IF) values of cytokines/chemokines/growth factors and used the blank subtracted IF values to compute z unit-based composite scores reflecting different immune profiles ^5,36,38,39^. ESF, Table 2 lists the immune profiles assessed in the current report.

**Table 1.**
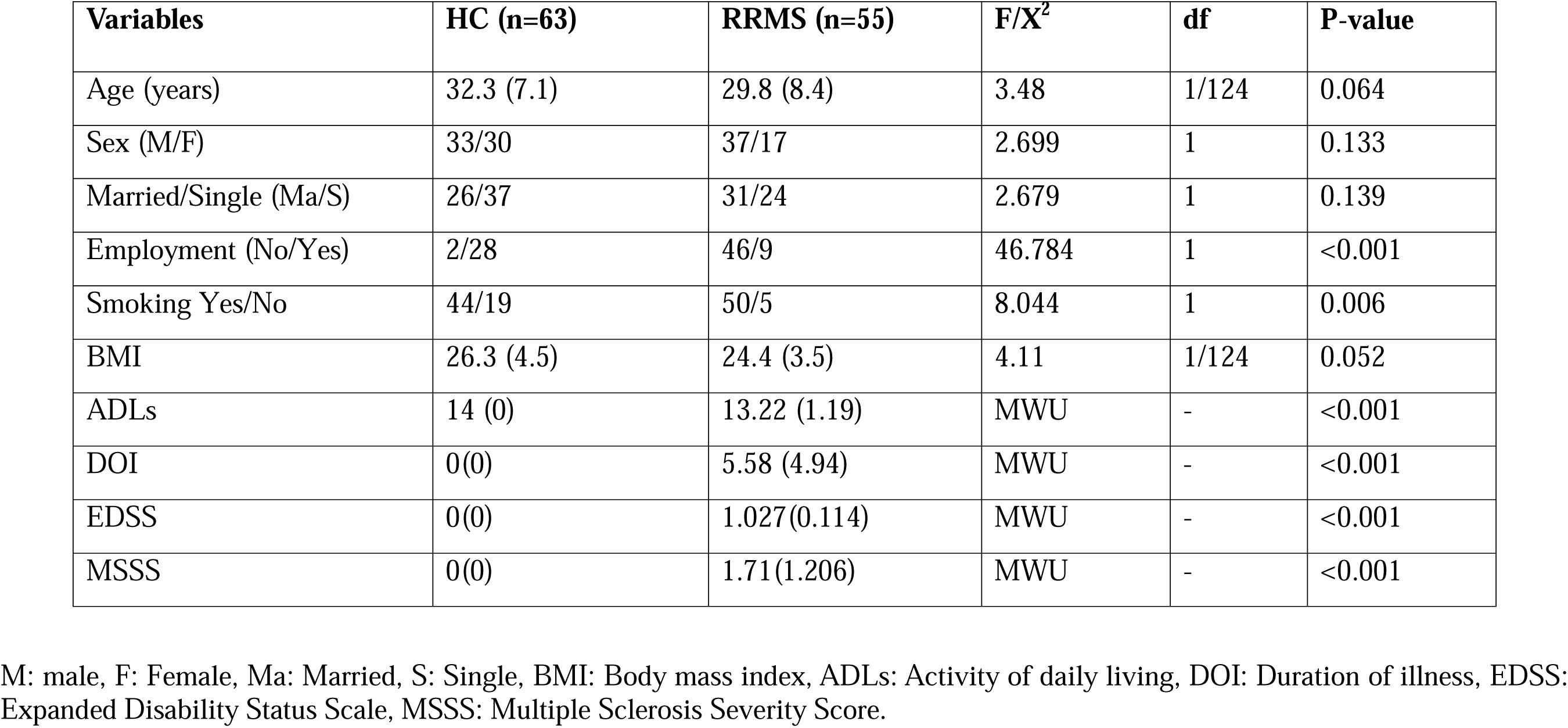
Demographic and clinical data in patients with relapsing remitting multiple sclerosis (RRMS) and healthy controls (HC).

| **Variables** | **HC (n=63)** | **RRMS (n=55)** | **F/X<sup>2</sup>** | **df** | **P-value** |
| --- | --- | --- | --- | --- | --- |
| Age (years) | 32.3 (7.1) | 29.8 (8.4) | 3.48 | 1/124 | 0.064 |
| Sex (M/F) | 33/30 | 37/17 | 2.699 | 1 | 0.133 |
| Married/Single (Ma/S) | 26/37 | 31/24 | 2.679 | 1 | 0.139 |
| Employment (No/Yes) | 2/28 | 46/9 | 46.784 | 1 | <0.001 |
| Smoking Yes/No | 44/19 | 50/5 | 8.044 | 1 | 0.006 |
| BMI | 26.3 (4.5) | 24.4 (3.5) | 4.11 | 1/124 | 0.052 |
| ADLs | 14 (0) | 13.22 (1.19) | MWU | - | <0.001 |
| DOI | 0(0) | 5.58 (4.94) | MWU | - | <0.001 |
| EDSS | 0(0) | 1.027(0.114) | MWU | - | <0.001 |
| MSSS | 0(0) | 1.71(1.206) | MWU | - | <0.001 |

M: male, F: Female, Ma: Married, S: Single, BMI: Body mass index, ADLs: Activity of daily living, DOI: Duration of illness, EDSS: Expanded Disability Status Scale, MSSS: Multiple Sclerosis Severity Score.

**Table 2.** Differences in immunoglobulin (Ig) IgG/IgA/IgM reactivity against Epstein–Barr virus (EBV) and Human herpesvirus type 6 (HHV-6) proteins and immune profiles between patients with relapsing remitting multiple sclerosis (RRMS) versus healthy controls (HC).

| **Variables** | **HCs (n=63)** | **RRMS (n=55)** | **F/X<sup>2</sup>** | **df** | **P** |
| --- | --- | --- | --- | --- | --- |
| IgG-EBNA-366-406 | 0.619(0.115) | 1.528(0.124) | 26.720 | 4/113 | <0.0001 |
| IgM-EBNA-366-406 | 0.298(0.029) | 0.580 (0.031) | 42.120 | 4/113 | <0.0001 |
| IgA-EBNA-366-406 | 0.290 (0.026) | 0.527(0.028) | 35.210 | 4/113 | <0.0001 |
| IgG-EBV-dUTPase | 0.836(0.098) | 1.927(0.105) | 53.964 | 4/113 | <0.0001 |
| IgM-EBV-dUTPase | 0.609(0.061) | 1.234(0.066) | 44.831 | 4/113 | <0.0001 |
| IgA-EBV-dUTPase | 0.511(0.076) | 1.367(0.081) | 55.131 | 4/113 | <0.0001 |
| IgG-HHV-6-dUTPase | 0.307(0.036) | 0.739(0.038) | 62.764 | 4/113 | <0.0001 |
| IgM-HHV-6-dUTPase | 0.326(0.032) | 0.694(0.035) | 55.459 | 4/113 | <0.0001 |
| IgA-HHV-6-dUTPase | 0.312(0.032) | 0.628(0.034) | 43.104 | 4/113 | <0.0001 |
| M1 macrophage (z scores) | -0.573(0.112) | 0.573(0.112) | 8.161 | 4/105 | 0.005 |
| Th1 (z scores) | -0.280(0.134) | 0.280(0.134) | 67.938 | 4/105 | <0.0001 |
| Th17 (z scores) | -0.652(0.108) | 0.652(0.108) | 53.600 | 4/105 | <0.0001 |
| Chemokines (z scores) | -0.607 (0.113) | 0.607(0.113) | 61.870 | 4/105 | <0.0001 |
| GFs (z scores) | -0.630(0.109) | 0.630(0.109) | 58.254 | 4/105 | <0.0001 |
| IRS (z scores) | -0.621(0.111) | 0.621(0.111) | 152.375 | 4/105 | <0.0001 |
| CIRS (z scores) | -0.790(0.087) | 0.790(0.087) | 152.438 | 4/105 | <0.0001 |
| IRS+CIRS (z scores) | -0.793(0.088) | 0.793(0.088) | 8.161 | 4/105 | 0.005 |

Ig: Immunoglobulin, EBNA: EBV nuclear antigen, dUTPase: deoxyuridine-triphosphatase, HHV-6: Human herpesvirus 6, EBV: Epstein–Barr virus, M1: Macrophage M1, Th: T helper, GFs: Growth factors, IRS: immune response system, CIRS: compensatory immune response system, IRS+CIRS: combined index of IRS and CIRS activation.

### Data analysis

We employed IBM’s SPSS 29 for all statistical analyses in this study. Analysis of variance (ANOVA) was used to compare continuous variables between study groups, while contingency table analysis was used to examine the association between categorical variables. Pearson’s product moment and point-biserial correlations were used to examine associations among scale variables and between scale and binary data, respectively. Multiple comparisons and associations were p-corrected for False-Discovery Rate (FDR). To explore the relationships between IgA, IgG, and IgM responses and RRMS diagnosis, we conducted a binary logistic regression analysis, using RRMS as dependent variable and healthy controls as the reference group. This analysis accounted for potential confounding factors such as age, gender, smoking, and BMI. Key results included estimates of effect sizes using Nagelkerke pseudo-R square, Wald statistics with p-values, odds ratios with 95% confidence intervals (CI), and B values (standard error, SE). For predicting outcomes related to the EDSS and the MSSS in RRMS patients, we utilized multivariate regression analysis, adjusting for demographic variables such as age, gender, and BMI. Manual and stepwise methods were applied, the latter with entry and exit criteria set at p-values of 0.05 and 0.06, respectively. This analysis provided model metrics, including standardized beta coefficients, degrees of freedom (df), p-values, R², and F-statistics. To examine heteroskedasticity, we used the White test and the modified Breusch-Pagan test, and we assessed collinearity by evaluating tolerance and the variance inflation factor. A two-tailed design was used for all tests, with a significant threshold of 0.05.

In addition, we developed multilayer perceptron neural network models to distinguish RRMS patients from healthy controls. These models included IgG/IgM/IgA responses to EBNA-366-406, EBV-DUTPase, HHV-6-DUTPase with or without immune profiles as input variables. The neural networks featured a feedforward architecture with two hidden layers, each potentially containing up to eight nodes, and they were trained in batch-type sessions for a maximum of 250 epochs. Training stopped when further reductions in the error term were not observed. The significance and relative importance of the input variables were visualized using an importance chart, and metrics such as error, relative error, and misclassification rates were calculated by comparing predicted versus actual values.

An a priori power analysis using G*Power 3.1.9.7 determined that a minimum sample size of 90 individuals was needed to detect differences in an ANCOVA (analysis of covariance) test, assuming a power of 0.8, a significance level (p) of 0.05, two groups, and 4 covariates and an effect size of 0.3.

## Results

### Socio-demographic and clinical characteristics of MS

**Table 1** presents the sociodemographic data, DOI, and scores from various disability assessments, including ADLs, the EDSS, and the MSSS, comparing RRMS patients with healthy controls. There were no significant differences observed in age, sex, marital status, or BMI between the groups. Significant differences were found between RRMS patients and healthy controls in employment status, and smoking status. RRMS patients demonstrated significantly elevated scores on ADL, EDSS, and MSSS scores.

### Differences in the immune responses among the study groups

**Table 2** shows that RRMS patients demonstrated higher IgG, IgA, and IgM responses to EBNA 366-406, EBV-dUTPase, and HHV-6-dUTPase as compared to healthy controls. In addition, M1 macrophage, Th17, chemokines, growth factor, IRS, CIRS, and IRS+CIRS profiles were significantly increased in MS patients.

In the restricted group of RRMS patients, there were no significant point-biserial correlations between the use of natalizumab and any of the viral data, except IgG-dUTPAse (r=-0.300, p=0.026, n=55, without false discovery rate p correction for multiple testing). The use of beta-interferon-1β was significantly associated (without p-correction) with lowered levels of IgM-EBNA (r=-0.283, p=0.036), IgA-EBNA (r=-0.336, p=0.012), IgM-EBV-dUTPase (r=–0.305, p=0.023), and IgA-HHV-6-dUTPase (r=-0.267, p=0.049). However, after using FDR p correction, all significances disappeared.

Binary logistic regression was used to identify the most important indicators for RRMS. In the logistic regression model, the control group functioned as the reference group and RRMS as the dependent variable. The IgA, IgG, and IgM responses to EBNA-366-406, EBV-dUTPase, and HHV-6-dUTPase, as well as age, sex, BMI, and smoking, were the independent factors in this investigation.

**Table 3**, model #1 indicates that IgG- and IgM-HHV-6-dUTPase were significantly and positively associated with RRMS with an effect size 0.803 and an overall accuracy of 91.5% (sensitivity=87.3%, and specificity=95.2%). Model #2 shows that these results did not change after including age, sex, BMI, and smoking.

**Table 3:** Results of binary logistic regression analysis with relapsing remitting multiple sclerosis (RRMS) as dependent variable.

| Dichotomies |  | Explanatory Variables | B | SE | Wald | P | OR | 95% CI |
| --- | --- | --- | --- | --- | --- | --- | --- | --- |
| **RRMS patients**<br>**versus healthy**<br>**controls** | **Model #1** | IgG-HHV-6-dUTPase | 4.622 | 1.043 | 19.621 | <0.001 | 101.68 | 13.15; 785.93 |
|  |  | IgM-HHV-6-dUTPase | 1.697 | 0.547 | 9.630 | 0.002 | 5.45 | 1.86; 15.93 |
|  | **Model #2** | IgG-HHV-6-dUTPase | 4.415 | 1.056 | 17.473 | <0.001 | 82.69 | 10.43; 655.48 |
|  |  | IgM-HHV-6-dUTPase | 1.734 | 0.572 | 9.178 | 0.002 | 5.66 | 1.84; 17.37 |

Ig: Immunoglobulin; DUTPase: deoxyuridine-triphosphatase, HHV-6: Human herpesvirus 6, BMI: Body mass index.
Model #1: Nagelkerke R square: 0.803; sensitivity: 95.2%; specificity: 87.3%; overall accuracy 91.5%.
Model #2: this model includes age, sex, BMI, and smoking (forced entry); these 4 variables were non-significant in the regression, and the impact of the HHV-6 markers on RRMS did not change after forced entry of these possible confounders.

**Table 4** shows the results of neural network analyses. NN#1 revealed the characteristics of a first neural network model that differentiates RRMS patients from healthy controls. This model was constructed using 2 hidden layers with 2 units in hidden layer 1, 2 in hidden layer 2, and 2 units in the output layer. The error term was significantly lower in the testing than in the training sample, whilst the percentage of incorrect classifications was similar between the three samples, indicating that the model was not overtrained. This solution was better than the logistic regression, with a predictive accuracy (computed in the holdout sample) of 97.7% (sensitivity=100% and specificity=96%) and an area ROC curve of 0.997. The topmost important biomarkers (**Figure 1**) were in descending order of importance: the growth factor profile, IgG-HHV-6-dUTPase, IRS+CIRS, and IgG- and IgM-EBV-dUTPases.

**Figure 1.**
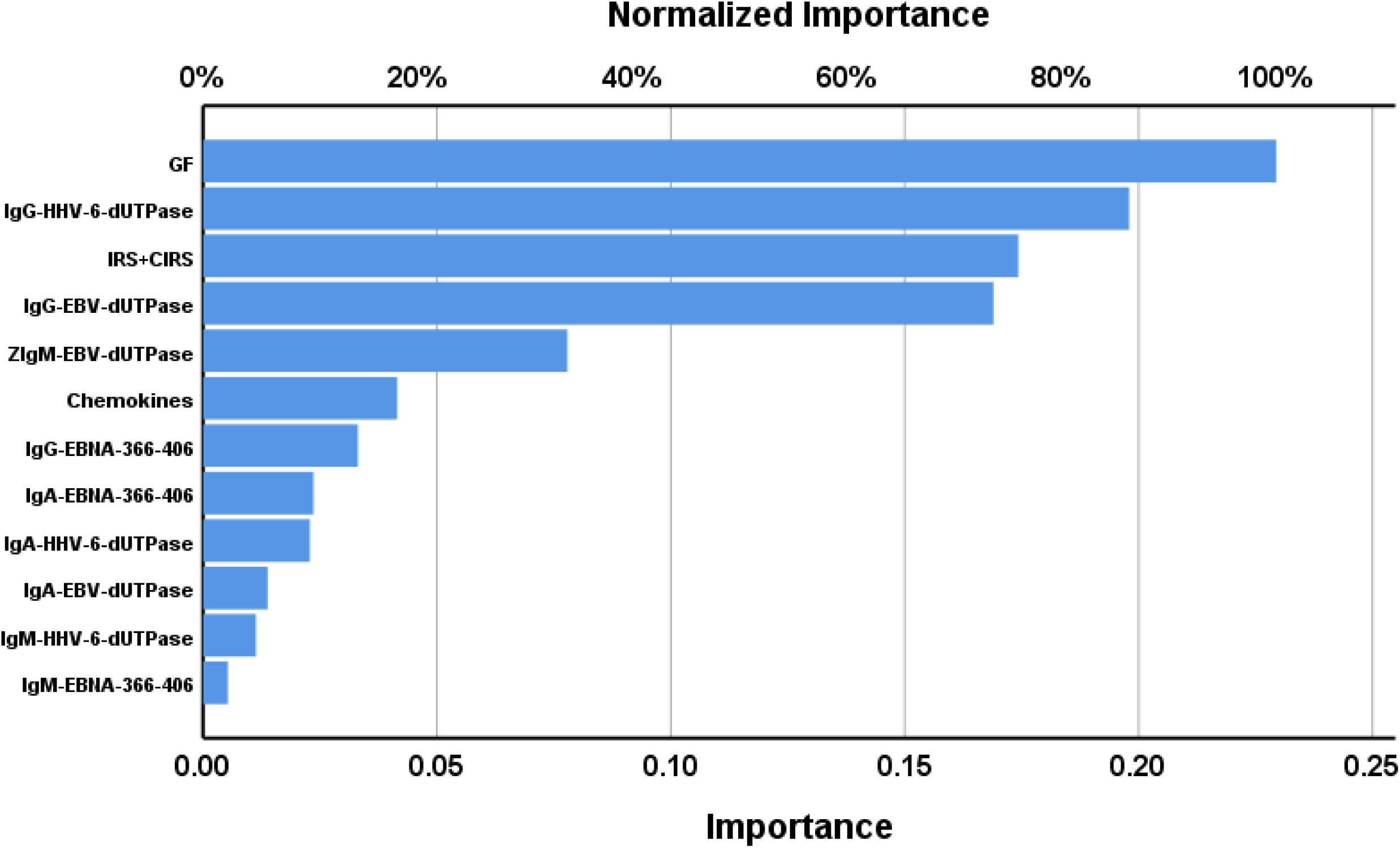
Results of neural network analysis demonstrating the importance chart. The output variables are the diagnosis of relapsing remitting multiple sclerosis and healthy controls. Input variables are growth factors (GF), Immunoglobulin (Ig)G-Human Herpes Virus 6 (HHV 6) deoxyuridine-triphosphatase (dUTPase), IRS+CIRS (immune activation index), IgG-Epstein–Barr virus (EBV) dUTPase, IgM-EBV-dUTPase, chemokines, IgG-EBV-nuclear antigen (EBNA)-366-406, IgA-EBNA-366-406, IgA-HHV6-dUTPase, IgA-EBV-dUTPase, IgM-HHV-6-dUTPase, IgM-EBNA-366-406.

**Table 4.**
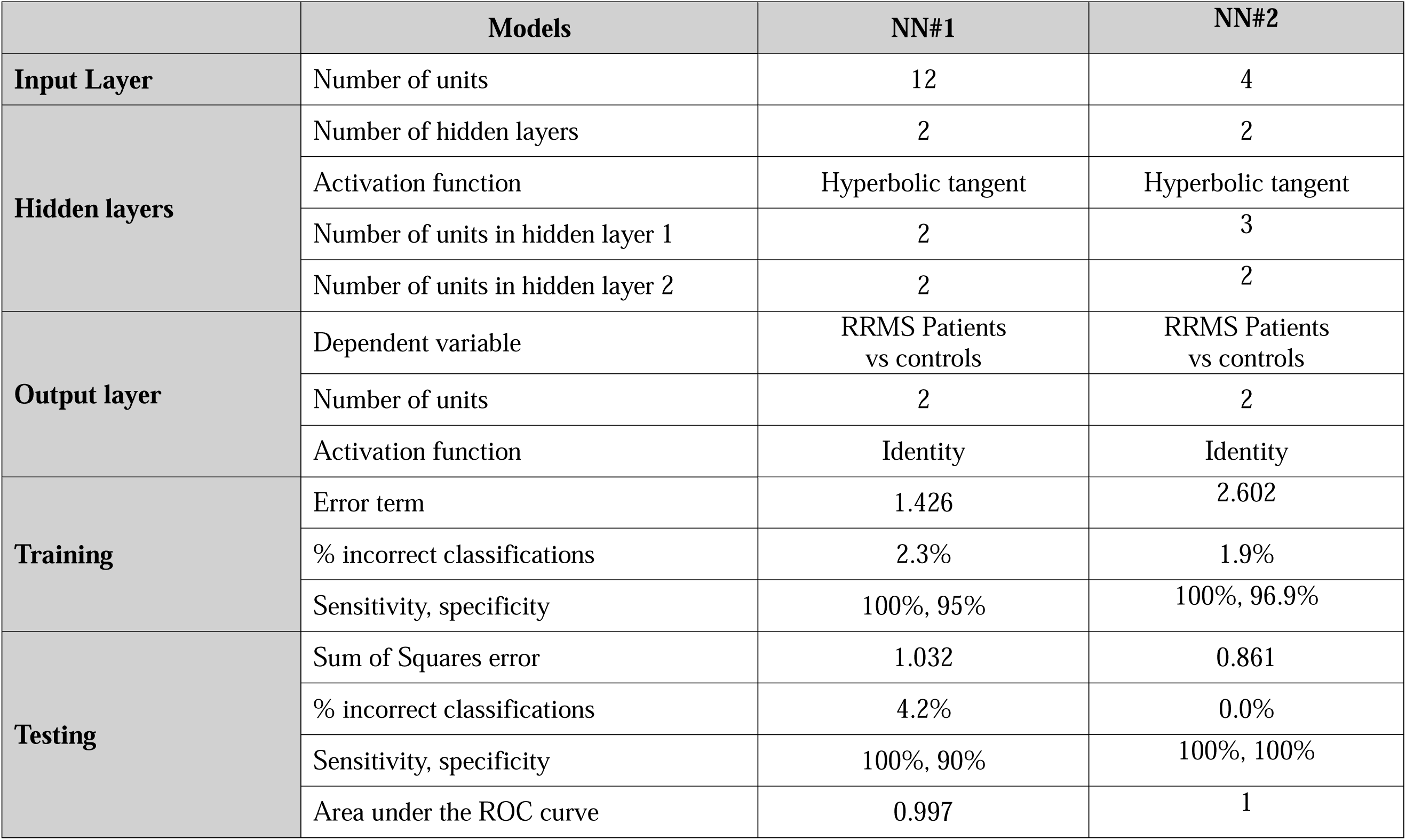
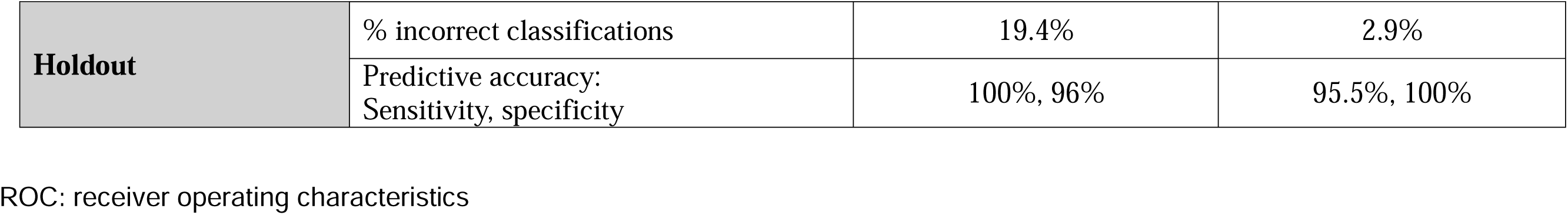
Results of neural network analysis with relapsing remitting multiple sclerosis (RRMS) as output variable.

We have rerun the above neural network analysis using the top 5 most important variables combined with a z unit composite score of IgG-EBV-dUTPase and IgG-HHV-6-dUTPase and detected that using 4 of those input variables yielded a comparable differentiation of RRMS from controls. The features of this second neural network model (NN#2) are displayed in **Table 4**. This model was constructed using 2 hidden layers with 3 nodes in hidden layer 1, 2 in hidden layer 2, and 2 units in the output layer. The error term was significantly lower in the testing than in the training sample, whilst the percentage of incorrect classifications was quite similar between the training, testing and holdout samples. The predictive accuracy of this neural network model (computed in the holdout sample) was 97.1% (sensitivity=95.5% and specificity=100%) with an area under the ROC curve of 1. The topmost important biomarkers (**Figure 2**) were in descending order of importance: IRS+CIRS, the composite of IgGs directed against dUTPases, growth factors, and IgM-EBV-dUTPase.

**Figure 2.**
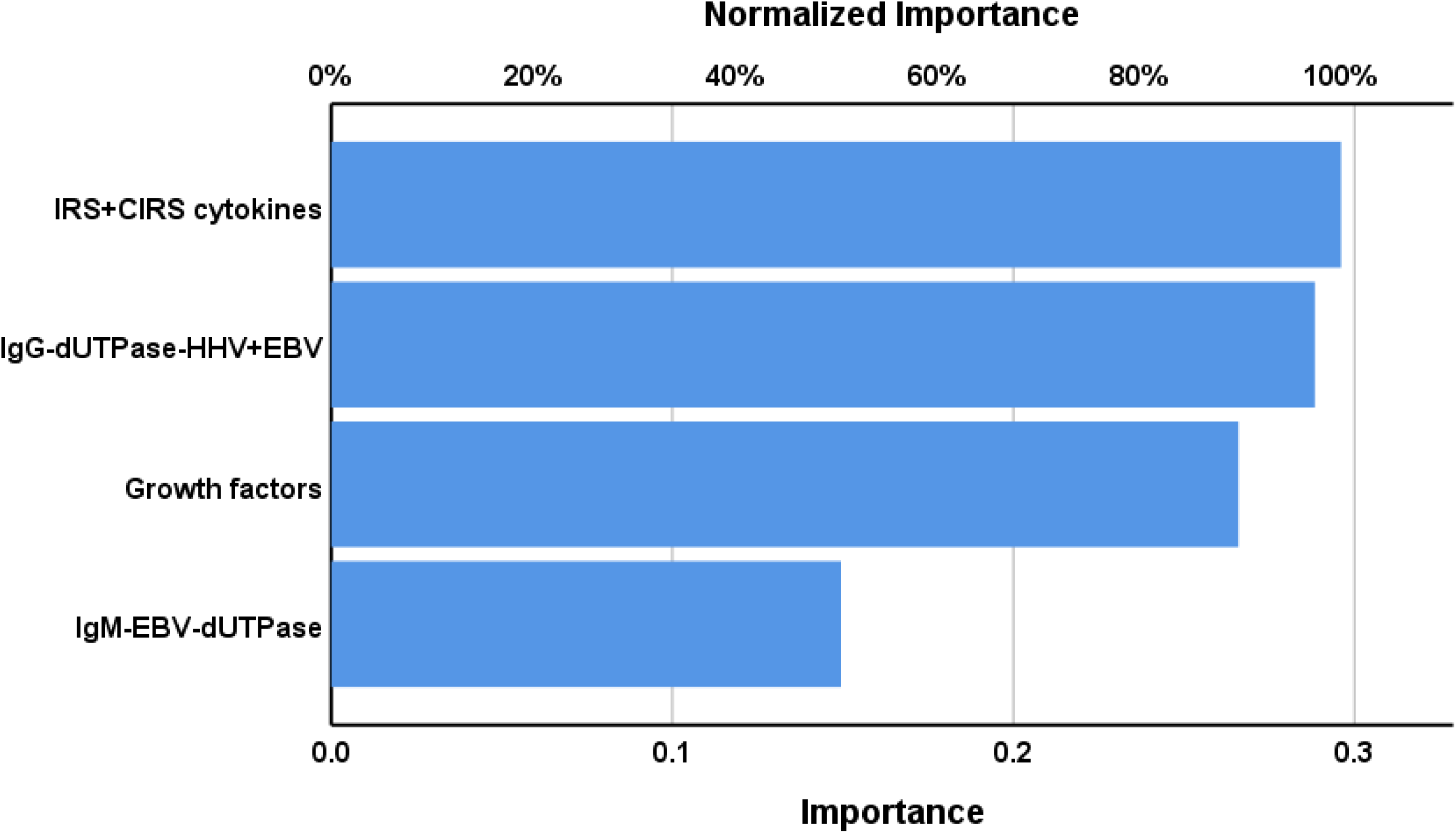
Results of neural network analysis demonstrating the importance chart. The output variables are the diagnosis of relapsing remitting multiple sclerosis and healthy controls. Input variables are IRS+CIRS (an immune activation index), a z unit composite score of IgG-EBV-dUTPase and IgG-HHV-6-dUTPase, growth factors, and IgM-EBV-dUTPase.

### Immune responses to reactivated latent viruses predict the severity of RRMS

**Table 5** presents the results of multiple regression analyses with the EDSS and MSSS scores as dependent variables. The independent variables include the IgG/IgM/IgA levels against the three viral antigens. First, we compute the effects of these viral data on disabilities (allowing for the effects of age, sex, smoking, and BMI). Consequently, we enter immune profiles into the multiple regression analyses. Regression analysis #1 shows that 65.4% of the variance in EDSS scores can be explained by IgG and IgM against HHV-6-dUTPase, and age. **Figure 3** illustrates the partial regression of the EDSS score on IgG against HHV-6-dUTPase. Regression analysis #2 shows that the model accounts for 77.6% of the variance in the EDSS score; IRS+CIRS, IgG and IgM against HHV-6-dUTPase, age, and sex were the significant explanatory variables. In regression analysis #3, IgG-HHV-6-dUTPase, IgM-EBV-dUTPase, and age predict 52.1% of the variance in MSSS scores. Regression #4 reveals that 64.4% of the variance in the MSSS score was explained by the regression on IgG-HHV-6-dUTPase, IRS+CIRS, IgA-HHV-6-dUTPase, and age.

**Figure 3:**
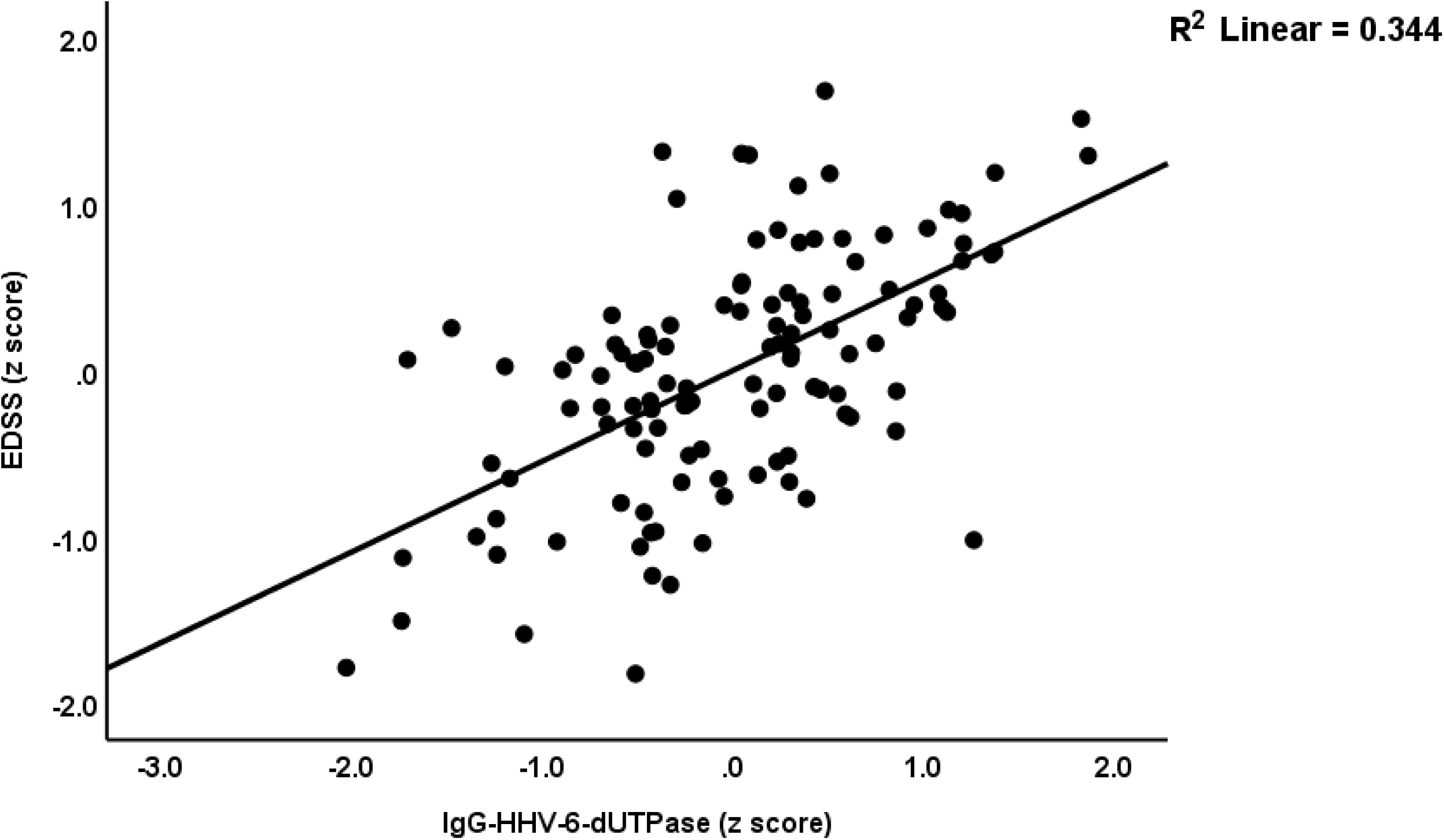
Partial regression of the Expanded Disability Status Scale (EDSS) on immunoglobulin (Ig)G directed against Human Herpes Virus-6 (HHV 6) deoxyuridine-triphosphatase (dUTPase), p < 0.001.

**Figure 4:**
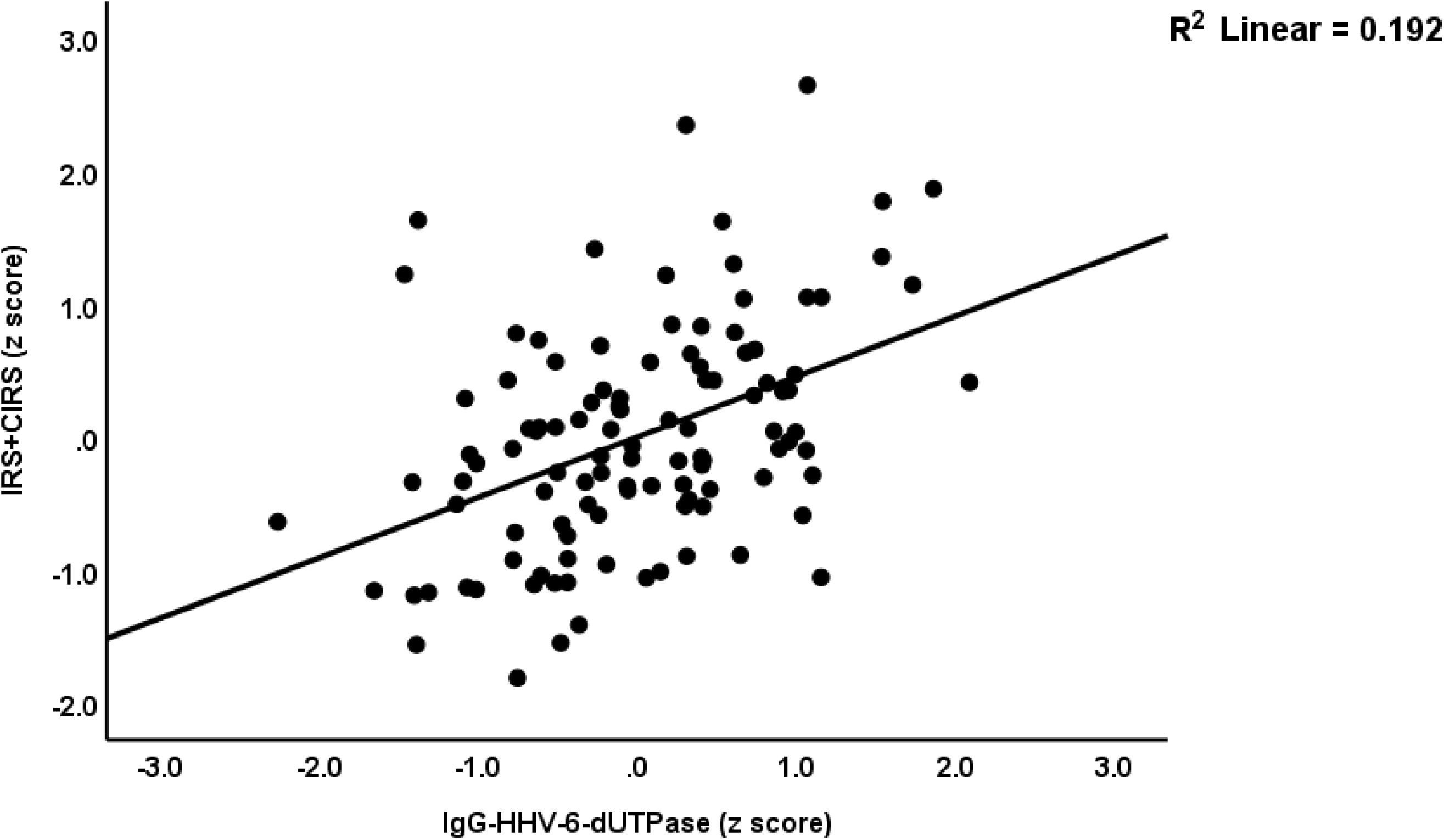
Partial regression of the immune activation index (IRS+CIRS) on immunoglobulin (Ig)G directed against Human Herpes Virus-6 (HHV 6) deoxyuridine-triphosphatase (dUTPase), p < 0.001.

**Table 5.**
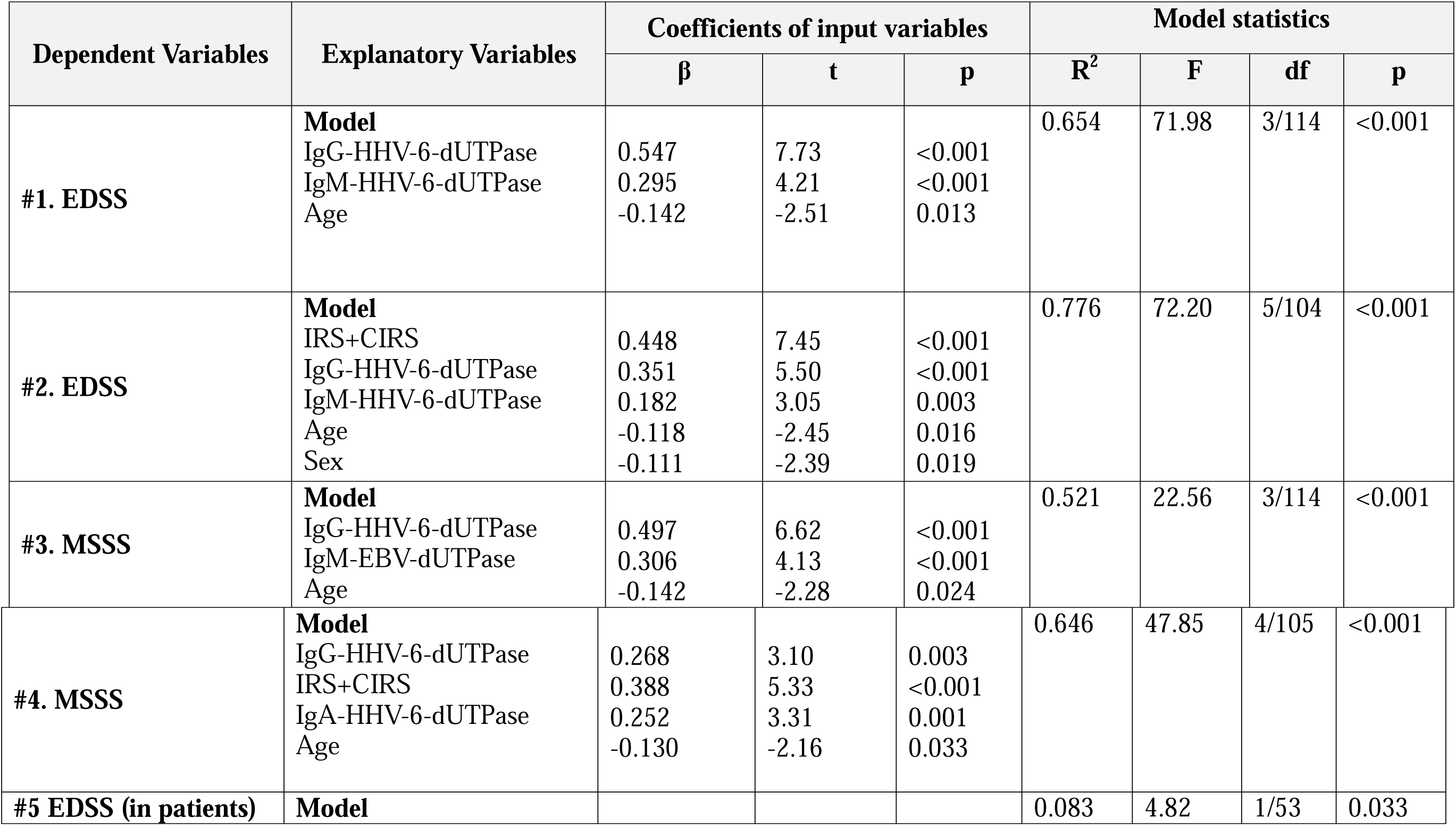
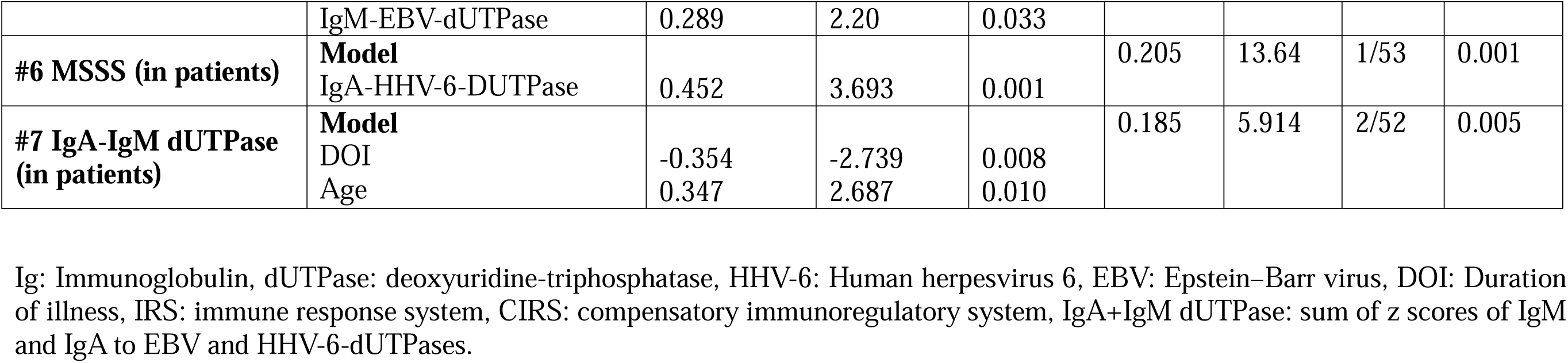
Results of multiple regression analysis with the Expanded Disability Status Scale (EDSS) and Multiple Sclerosis Severity Score (MSSS) as dependent variables.

We conducted additional regression analyses (#5 and #6), which were performed in the restricted study group of RRMS patients. Regression analysis #5 shows that 8.3% of the variance in EDSS score is explained by the regression on IgM-EBV-dUTPase (positively associated with the outcome). IgA-HHV-6-dUTPase accounted for 20.5% of the variance in the MSSS score (regression #6). Lastly, we also examined whether the IgA/IgM values to the dUTPases were predicted by DOI. Toward this end we computed a z composite score using the IgA/IgM responses to EBV and HHV-6-dUTPases. We found that this composite score was inversely associated with DOI and positively with age. Both variables together explained 18.5% of the variance of the composite score.

### Reactivated latent viruses and immune profiles

**Table 6** shows the associations between the immune profiles and the EBV and HHV-6 data. The latter were entered as explanatory variables, whereas the immune profiles were entered as dependent variables. In addition, we allowed for the effects of age, sex, BMI, and smoking. Regression #1 shows that the model accounts for 17.3% of the variance in the M1 macrophage profile, with IgG-HHV-6-dUTPase and IgM-EBV-dUTPase emerging as significant predictors. Regression #2 shows that these two variables explained 27.2% of the variance in the Th17 profile. Th1 was not predicted by any of the variables. Regression #3 demonstrates that 21.8% of the variance in GFs is significantly explained by IgG-HHV-6-dUTPase and IgM-EBV-dUTPase, both of which are positively associated with the Th17 axis. In regression #4, 16.3% of the variance in IRS is significantly explained by IgG-HHV-6-dUTPase. Regression #5 shows that 52.3% of the variance in CIRS is significantly explained by IgG-HHV-6-dUTPase, IgM-EBNA-366-406, and smoking. Regression #6 reveals that 40.7% of the variance in the IRS+CIRS ratio is explained by IgG-HHV-6-dUTPase and IgM-EBV-dUTPase. Regression #7 indicates that IgG-HHV-6-dUTPase explains 15.8% of the variance in the chemokine profile.

**Table 6.** Results of multiple regression with the immune profiles as dependent variables.

| Dependent Variables | Explanatory Variables | Coefficients of input variables |  |  | Model statistics |  |  |  |
| --- | --- | --- | --- | --- | --- | --- | --- | --- |
| | | $\beta$ | t | p | R<sup>2</sup> | F | df | p |
| #1. M1 macrophage | **Model** |  |  |  | 0.173 | 11.22 | 2/107 | <0.001 |
|  | IgG-HHV-6-dUTPase | 0.258 | 2.458 | 0.016 |  |  |  |  |
|  | IgM-EBV-dUTPase | 0.215 | 2.051 | 0.043 |  |  |  |  |
| #2. Th17 | **Model** |  |  |  | 0.272 | 19.94 | 2/107 | <0.001 |
|  | IgG-HHV-6-dUTPase | 0.380 | 3.858 | <0.001 |  |  |  |  |
|  | IgM-EBV-dUTPase | 0.205 | 2.082 | 0.040 |  |  |  |  |
| #3. GFs | **Model** |  |  |  | 0.218 | 14.88 | 2/107 | <0.001 |
|  | IgG-HHV-6-dUTPase | 0.296 | 2.903 | 0.004 |  |  |  |  |
|  | IgM-EBV-dUTPase | 0.233 | 2.284 | 0.024 |  |  |  |  |
| #4. IRS | **Model** |  |  |  | 0.163 | 21.04 | 1/108 | <0.001 |
|  | IgG-HHV-6-dUTPase | 0.404 | 4.587 | <0.001 |  |  |  |  |
| #5 CIRS | **Model** |  |  |  | 0.523 | 38.76 | 3/106 | <0.001 |
|  | IgG-HHV-6-dUTPase | 0.432 | 5.166 | <0.001 |  |  |  |  |
|  | IgM-EBNA-366-406 | 0.303 | 3.778 | <0.001 |  |  |  |  |
|  | Smoking Y/N | -0.163 | -2.252 | 0.026 |  |  |  |  |
| #6 IRS+CIRS | **Model** |  |  |  | 0.407 | 36.74 | 2/107 | <0.001 |
|  | IgG-HHV-6-dUTPase | 0.448 | 5.039 | <0.001 |  |  |  |  |
|  | IgM-EBV-dUTPase | 0.272 | 3.056 | 0.003 |  |  |  |  |
| #7 Chemokines | **Model** |  |  |  | 0.158 | 20.29 | 1/108 | <0.001 |
|  | IgG-HHV-6-dUTPase | 0.398 | 4.505 | <0.001 |  |  |  |  |

Ig: Immunoglobulin, dUTPase: deoxyuridine-triphosphatase, HHV-6: Human herpesvirus type 6, EBV: Epstein–Barr virus. M1: Macrophage M1, Th: T helper, GFs: Growth factors, IRS: Immune response system, CIRS: Compensatory immunoregulatory system, IRS+CIRS: combined index of IRS and CIRS activation.

## Discussion

### RRMS is associated with EBV and HHV-6 reactivation

The first major finding of this study is that remitted RRMS patients demonstrated significantly increased levels of IgG/IgA/IgM to HHV-6 and EBV viral proteins, including EBNA-366-406, EBV-dUTPase, and HHV-6-dUTPase as compared to normal controls. Moreover, our logistic regression analysis shows that a combination of the IgG and IgM directed to HHV-6 dUTpase may successfully separate RRMS from healthy controls with an overall accuracy of 91.5%.

The detection of antibody responses to HHV-6 in the CSF, or the presence of HHV-6 DNA in serum, CSF supernatant, or CSF leukocytes, is typically uncommon during clinical remission of MS, highlighting the limitations of these markers for a definitive diagnosis ^40^. Nevertheless, the measurement of IgG to dUTPase (and by inference IgM and IgA to the dUTPase), are more specific markers of ongoing viral replication. Elevated IgG levels against herpesvirus dUTPases suggest ongoing viral replication through an abortive lytic cycle, where viral proteins are produced without forming new virions ^41,42^. It should be stressed that this kind of replication markers does not necessarily indicate increased viral load ^41^.

The current study extends previous evidence which reported elevated IgG responses to EBV EBNA-1 and HHV-6 in MS patients ^22,23^. Our results extend those of previous studies which reported increased antibodies (IgM and IgG) to HHV-6 and EBV ^19,24,43^. A recent meta-analysis reported an association between MS and EBV and HHV-6^18^. In addition, previous studies have suggested an association between HHV-6 and MS based on immunological and molecular data, indicating that a subset of MS patients may show HHV-6 reactivation ^44–46^.

Importantly, we also observed that a combination of IgG, IgM, and IgA responses to HHV-6 and EBV-dUTPase significantly predicted the severity of disabilities due to MS (as measured by the EDSS) and disease progression (as assessed with the MSSS scale). These findings extend previous research that linked IgG against HHV-6 with an increased risk of MS progression, including relapse and disability ^43,47^. Lundström and Gustafsson reported that HHV-6A infection may play a role in the early stages of MS ^21^. Anti-HHV-6A/B IgG titers also hold the potential to predict impending relapses in MS disease ^48^. The presence of HHV-6 DNA in serum samples of MS patients coupled with increased IgG response to HHV-6 suggests a possible association between virus reactivation and disease progression ^45^. However, other studies contradict these results by reporting that the parameters of EBV and HHV-6 reactivation were not always associated with disability or progression of MS disease ^49,50^.

Overall, the findings of the present study indicate a significant role of EBV and HHV-6 replication in the pathogenesis of RRMS and its disabilities and progression. Both increased EBV and HHV-6 replication may, through different mechanisms, induce MS relapses. HHV-6 reactivation may trigger or aggravate autoimmunity and tissue damage associated with MS lesion development, possibly through molecular mimicry or excessive complement activation ^44^. Lanz et al. reported a high-affinity molecular mimicry between the EBNA1 and the CNS protein GlialCAM in MS patients ^51^. Clonally expanded B cells in the CSF of MS patients produce antibodies that cross-react with EBNA1 and GlialCAM, facilitated by post-translational modifications. This cross-reactivity was shown to exacerbate disease in a mouse model of MS, providing a mechanistic link between EBV infection and MS pathobiology ^51^. Serum levels of the neurofilament light chain, a biomarker of neuroaxonal degeneration, increased only following EBV seroconversion, indicating a temporal relationship between EBV infection and the onset of MS. This suggests that EBV plays a crucial role in the pathogenesis of MS and highlights the potential for preventive strategies targeting EBV to reduce MS incidence ^52^.

Interestingly, we found that the IgA responses to EBV and HHV-6 dUTPases were inversely correlated with the duration of illness. These findings may suggest that the replication ability of these viruses decreases with illness duration. Such an effect could play a role in the age-associated decreases in annualized relapse rate ^53^.

### Viral replication and the drug state of the RRMS patients

In the current study, the use of disease-modifying therapies (DMTs) had, after FDR p correction, no significant effects on the EBV and HHV-6 antibody data. We found a trend toward a suppressant effect of beta-interferon-1β on the dUTPase values. Beta-interferon-1β treatment has been reported to decrease HHV-6 viral load during MS relapses, though this effect is not observed during remission ^54^. Treatment with Natalizumab is associated with the reactivation of several latent viruses, including HHV-6 ^55^ and EBV ^56,57^. This reactivation is believed to result from impaired immune surveillance, a consequence of Natalizumab’s impact on T-cell migration and phenotype ^55,58^. Moreover, the measurement of anti-HHV-6 IgG titers has been proposed as a potential biomarker for the clinical response to DMTs ^48^. High IgG response to EBNA-1 correlated with MS disease activity on MRI and disability progression along with clinical response to natalizumab ^25,59^.

Overall, our results indicate that even in the remitted phase of RRMS and despite treatments with DMTs, HHV-6 and EBV replication are present and, consequently, may contribute to new relapses.

### EBV and HHV-6 antibodies and immune profiles

Almulla et al. (2023) demonstrated that elevated levels of Th-1 and Th-17 cytokines, are significant predictors of disabilities in MS patients ^5^. This finding is particularly relevant given that the upregulation of these immune profiles may contribute to autoimmune processes through diverse mechanisms ^60^.

Most importantly, the current report shows that the EBV and HHV-6 replication indices are significantly correlated with activated immune profiles, including overall immune functions (IRS+CIRS), M1, Th-17, IRS, CIRS and growth factor immune profiles. For example, around 20% of the variance in the M1 and Th-17 profiles was explained by the combined effects of IgM or IgG directed to EBV-dUTPase and HHV-6-dUTPase. While 16.3% of the variance in the IRS profile was associated with IgG HHV-6-dUTPase, 40% of the variance in CIRS profile was associated with both EBV (IgM) and HHV-6 (IgG)-dUTPases. Thus, it appears that dUTPases of both EBV and HHV-6 may drive IRS and CIRS components.

dUTPases are recognized as pathogen-associated molecular pattern proteins that may exacerbate immune pathology in various diseases including MS ^29^. These viral proteins could influence disease pathophysiology by modulating the host innate immune system specifically by triggering cytokine production (M1, Th-1 and Treg cytokines), and activating Toll-Like receptors and NF-κB ^61–63^. In addition, EBV-dUTPase contributes to latency by increasing the production of IL-21 and activin A ^42^. HHV-6 seropositivity was linked to altered pro-inflammatory cytokine levels in MS patients ^64^. The correlation between HHV-6 reactivation and serum IL-12 concentrations during disease activity suggests that HHV-6 reactivation is linked to MS aggravation ^65^.

Furthermore, the current study provides evidence that combining EBV and HHV-6 replication indices with growth factor and IRS+CIRS immune profiles yields a predictive accuracy of 97.1% in diagnosing RRMS. Thus, EBV and HHV-6 replication and its consequences, i.e., immune activation, may be used to externally validate the diagnosis of RRMS. Previous research has focused on other biomarkers such as neurofilament light chain, chitinase 3-like proteins, and microRNAs ^66,67^. The combination of several CSF and plasma proteins, such as IL-12B, CD5, MIP-1a, and CXCL9, had a diagnostic efficacy similar to traditional markers, including IgG index and neurofilament light chain ^68^. In addition, serum biomarkers, including adhesion molecules and matrix metalloproteinase-9, may help monitor disease activity ^69^.

## Limitations

When interpreting the results of this study, certain limitations must be acknowledged. Considering the role of oxidative and nitrosative stress markers in the pathophysiology and progression of MS ^70,71^, it would be beneficial to examine these biomarkers in conjunction with indicators of latent viral reactivation.

## Conclusions

IgG/IgA/IgM responses to EBV and HHV-6 dUTPase combined with immune profiles allow to externally validate the diagnosis of RRMS. EBV and HHV-6 replication are associated with disabilities and progression of the disease. Our results suggest that the pathophysiology of MS involves both the reactivation/replication of latent viruses and the activation of immune-inflammatory pathways. Different mechanisms related to EBV and HHV-6 replication may explain the onset and relapsing of MS, including inflammation, expansion of M1 macrophages and Th-17 cells, molecular mimicry, infiltration of autoreactive T cells and antibodies, neuroinflammation, and breakdown of myelin sheaths. Our findings highlight that EBV and HHV-6 dUTPases contribute to these pathways and indicate that EBV/HHV-6 dUTPases are new drug targets to treat RRMS and prevent future relapses.

## Data Availability

AA managed the blood sample collection and patient-related procedures. AV and AFA conducted the serum biomarker quantification. MM performed the statistical analysis. MM, AV, and EV performed visualization. The initial draft was written by AA and MM, and subsequently revised by AV, YZ, and EV. All authors approved of the last version.

## Acknowledgements

The authors extend their sincere appreciation to the Neuroscience Center of Alsader Medical City in Al-Najaf province, Iraq, for their crucial assistance in gathering the data.

## Ethical approval and consent to participate

The investigation received approval from the Ethics Committee of the College of Medical Technology at the Islamic University of Najaf, Iraq (Document No. 11/2021). All procedures adhered to both Iraqi and international ethical standards and written informed consent was obtained from all patients and control participants.

## Declaration of interest

The authors declare no conflicts of interest.

## Funding

Funding for the project was provided by the C2F program at Chulalongkorn University in Thailand, grant number 64.310/436/2565 to AFA, the Thailand Science Research, and Innovation Fund at Chulalongkorn University (HEA663000016), and a Sompoch Endowment Fund (Faculty of Medicine) MDCU (RA66/016) to MM. For performance of all antibody assays, funds were provided by Immunosciences Lab., Inc., Los Angeles, CA, USA, and Cyrex Labs, LLC, Phoenix, AZ, USA.

## Availability of data

Upon receiving a valid request and following a thorough analysis of the data, the corresponding author (MM) is prepared to provide access to the SPSS file associated with this study.

## Notes

### Competing Interest Statement

The authors have declared no competing interest.

