## supplementary file for "Reactivation of Human Herpesvirus 6 and Epstein-Barr Virus in relapsing remitting multiple sclerosis: association with disabilities, disease progression, and inflammatory processes"

**ELECTRONIC SUPPLEMENTARY FILE (ESF)**

**ESF, Table 1.** Cytokine, chemokines and growth factors examined in the current study

| **Protein abbreviations** | Gene Symbol | **Protein name** |
| --- | --- | --- |
| IL-1β | IL1B | Interleukin-1β |
| IL-1RA | IL1RN | Interleukin-1 receptor antagonist |
| IL-2 | IL2 | Interleukin-2 |
| IL-4 | IL4 | Interleukin-4 |
| IL-5 | IL5 | Interleukin-5 |
| IL-6 | IL6 | Interleukin-6 |
| IL-7 | IL7 | Interleukin-7 |
| CXCL8 | CXCL8 | C-X-C motif chemokine ligand 8 (IL-8) |
| IL-9 | IL9 | Interleukin-9 |
| IL-10 | IL10 | Interleukin-10 |
| IL-12p70 | IL12 | Interleukin-12 |
| IL-13 | IL13 | Interleukin-13 |
| IL-15 | IL15 | Interleukin-15 |
| IL-17 | IL17 | Interleukin-17 |
| CCL11 | CCL11 | Eotaxin |
| FGF2 | FGF2 | Fibroblast growth factor 2, Basic fibroblast growth factor |
| G-CSF | CSF3 | Granulocyte Colony Stimulating Factor, Colony Stimulating Factor 3 (Granulocyte) |
| GM-CSF | CSF2 | Granulocyte-macrophage colony-stimulating factor, Colony-stimulating factor 2 |
| IFN-γ | IFNG | Interferon-γ |
| CXCL10 | CXCL10 | C-X-C motif chemokine ligand 8, Interferon gamma-induced protein 10 (IP10) |
| CCL2 | CCL2 | C-C Motif Chemokine Ligand 2 (MCP1) |
| CCL3 / MIP-1α | CCL3 | Macrophage inflammatory protein-1 alpha, C-C Motif Chemokine Ligand 3 |
| PDGF | PDGFA | Platelet Derived Growth Factor Subunit A |
| CCL4 / MIP-1β | CCL4 | C-C Motif Chemokine Ligand 4, Macrophage Inflammatory Protein 1-Beta, Lymphocyte Activation Gene 1 Protein |
| CCL5 /RANTES | CCL5 | C-C Motif Chemokine Ligand 5, Regulated Upon Activation, Normally T-Expressed, And Presumably Secreted |
| TNF-α | TNF | Tumor Necrosis Factor-Alpha |
| VEGF | VEGFA | Vascular Endothelial Growth Factor |

**ESF, Table 2**. Description of the immune profiles used in this study

| **Immune Profile** | **Members** |
| --- | --- |
| **M1 macrophage** | IL-1β, sIL-1RA, IL-6, TNF-α, CXCL8, CCL3 |
| **T helper-1** | IL-2, IFN-γ, IL-12 |
| **T helper-17** | IL-6, IL-17 |
| **IRS** | IL-1β, IL-6, TNF-α, CXCL8, CCL3, IL-2, IFN-γ, IL-12, IL-17, IL-15, G-CSF, GM-CSF, CXCL10, CCL5, CCL2 |
| **CIRS** | IL-4, IL-10, sIL-1RA |
| **IRS+CIRS** | IL-1β, IL-6, TNF-α, CXCL8, CCL3, IL-2, IFN-γ, IL-12, IL-17, IL-15, G-CSF, GM-CSF, CXCL10, CCL5, CCL2, IL-4, IL-10, sIL-1RA |
| **Chemokines** | CXCL8, CCL11, CXCL10, CCL2, CCL3, CCL4, CCL5 |
| **Growth factors** | FGF, PDGF, VEGF |

IRS: immune-inflammatory response system; CIRS: compensatory immunoregulatory system
